# Utilization of wearable activity monitors to support physical activity interventions in neurodegenerative diseases: a feasibility study

**DOI:** 10.1101/2022.05.31.22275824

**Authors:** Hai-Jung Steffi Shih, Lori Quinn, Philippa Morgan-Jones, Katrina Long, Abigail R. Schreier, Ciaran P. Friel

**Affiliations:** Department of Biobehavioral Sciences, Teachers College, Columbia University, New York, NY; Department of Rehabilitation and Regenerative Medicine (Physical Therapy), Columbia University Irving Medical Center, New York, NY; School of Engineering, Cardiff University, UK; Centre for Trials Research, Cardiff University, UK; Department of Occupational Therapy, San Jose State University, San Jose, CA; Institute of Health System Science, Feinstein Institutes for Medical Research, Northwell Health, New York, NY

**Keywords:** Exercise, Wearable Sensors, Digital Health, Parkinson Disease, Huntington Disease, Coaching, Physical Therapy, Occupational Therapy

## Abstract

**Purpose:** To evaluate the feasibility of using wearable activity monitors as part of a physical activity (PA) intervention and describe patterns of device weartime and PA behavior in people with Parkinson (PD) and Huntington disease (HD).

**Materials and Methods:** Secondary analyses were conducted on two pilot studies where people with early-stage PD (n=13) and HD (n=14) enrolled in a 4-month PA coaching program. Participants wore a Fitbit Charge 2 and physical and occupational therapists guided them through understanding of PA data and goal-setting to build autonomy and facilitate PA uptake. Wear time, wear habits (delineated based on nighttime wear), and activity metrics including steps, resting heart rate, and metabolic equivalent of task (MET) *minutes were analyzed.

**Results:** Retention rate of the intervention was 85%. Participants had a mean (SD) of 92.3(9.2)% valid wear days over the intervention period. Average daily wear time was 18.4(4.5) hours, with an initial 2-week period of high wear time fluctuation. Regardless of diagnosis, day & night Fitbit wearers had more improvements in steps (d=1.02) and MET*min/week (d=0.69) compared to day-only wearers.

**Conclusions:** Implementing wearable activity monitors in a PA coaching intervention was feasible and provided insight into longitudinal PA behavior in people with neurodegenerative diseases.

## Introduction

Physical activity (PA) is essential for the maintenance of physical and mental wellbeing, and federal guidelines provide clear direction on frequency, intensity, and duration of exercise for most individuals.^1–3^ In modern-day life, it is arguably difficult for the general population to reach recommended levels of PA, let alone those with neurodegenerative diseases who have motor and cognitive impairments that may impact mobility.^4–6^ Insufficient PA is especially detrimental for people living with neurodegenerative diseases, such as Parkinson disease (PD) and Huntington disease (HD), as there is evidence that PA can slow functional decline and is potentially neuroprotective.^7–10^ PD and HD are neurodegenerative diseases that primarily affect the basal ganglia, resulting in movement disorders that over time become progressively more disabling.^11,12^ Due to the length of time over which neural degeneration occurs in both diseases, developing interventions that facilitate initiation of, and adherence to, regular PA early in the disease is critical and can set the stage for long-term disease self-management.^13,14^

Physical and occupational therapists are uniquely trained to provide consultation for PA uptake as they are trusted by patients and well-versed in exercise prescription, neurological disease presentation, and motivational interviewing. However, therapists are challenged by limited sessions of therapy and inadequate means of monitoring exercise outside of therapy.^15,16^ Remote monitoring devices such as wearable activity monitors can provide objective feedback on PA levels and have the potential to augment physical and occupational therapy interventions.^17–19^ Combining therapists’ expert consultation and wearable activity monitors may result in increased and sustained PA uptake.

Wearable activity monitors such as Fitbit, Garmin, and Apple Watch are now ubiquitous in daily life. There is growing research that incorporates these wearable devices, especially in fields focused on cancer, cardiovascular, and metabolic diseases.^20–23^ People with neurodegenerative diseases have unique barriers related to their disease presentation, specifically in terms of motor, cognitive, and mood function.^24–27^ Several studies have shown that wearable activity monitors may provide useful information that can be used to support PA uptake in people with neurodegenerative diseases.^28–30^ Beyond the potential of wearable activity monitor data as an outcome measurement, they are also valuable as a behavior change intervention tool.^31^ Wearable activity monitors can provide insight into individual PA levels and enable more individualized goal setting and monitoring.

This paper reports secondary analysis of two related pilot studies in which a commercially available PA monitor, Fitbit Charge 2, was used as part of a 4-month PA coaching intervention in people with PD and HD. The purpose of this paper is to: 1) evaluate the feasibility of Fitbit devices as part of a PA intervention in neurodegenerative diseases, 2) describe Fitbit wear patterns and PA behavior during the intervention, and 3) explore if Fitbit wear habits impacted improvement of PA levels during the intervention.

## Methods

### Coaching Intervention

*PreActive-PD* and *PreActive-HD* were two pilot studies of therapist-delivered coaching interventions designed for people with PD (NCT03696589) and HD (NCT03306888), respectively;^32^ both pilot studies were used to inform larger clinical trials (NCT03344601 & NCT05308238). The interventions were grounded in self-determination theory and coaching elements focused on promoting autonomy, competence, and relatedness to improve PA behavior.^33,34^ Logic models that describe components of these interventions are previously published.^35,36^ Participants received between 4 and 6 one-on-one therapist coaching sessions either in person or via telehealth over the course of 4 months. A physical or occupational therapist worked collaboratively with the participant using motivational interviewing^37^ skills while offering support and education. Disease-specific workbooks were provided to participants to reinforce the coaching sessions and self-monitoring.* Therapists provided structured support for goal-setting and evidence-based exercise options. The focus of the intervention was to increase levels of moderate to vigorous PA. To support autonomy, participants were encouraged to select activities that they deemed feasible and meaningful. Therapists provided feedback and strategies to overcome exercise barriers and supported planning for goal attainment.

### Fitbit Use

To track PA during the intervention, participants were offered the option of using a Fitbit or a written log, and all participants chose to use the Fitbit. Fitbit use served two main functions: 1) a tool for participants to self-monitor their activity, and 2) data for therapists to review so as to inform coaching. The Fitbits were given to participants in the first coaching session, and therapists assisted with setting up the user accounts. Heart rate maximum was based on age-predicted values as determined by Fitbit (220-age formula^38^). Participants were provided a ‘Fitbit 101 guide’ for basic functions such as how to sync and charge the device and access activity data on their smartphone or computer. Participants were instructed to wear the device as much as possible during the waking hours, but no specific instructions for night time use were given. The Fitbit accounts were synchronized to the Fitabase (Small Steps Labs LLC, San Diego, CA) platform, a secure cloud-based research platform that enables remote monitoring by the therapists. Prior to coaching sessions, therapists reviewed participants’ data since their prior session including daily steps, heart rate, and activity intensity. Fitabase provided graphic visualization of the aforementioned measures that helped facilitate discussion during the coaching sessions (figure 1). Heart rate data provided information on exercise intensity and duration that was helpful for participant’s understanding of effective exercise sessions. Exercise goals were reviewed and revised based on wearable data and individual factors.^39^

**Figure 1.**
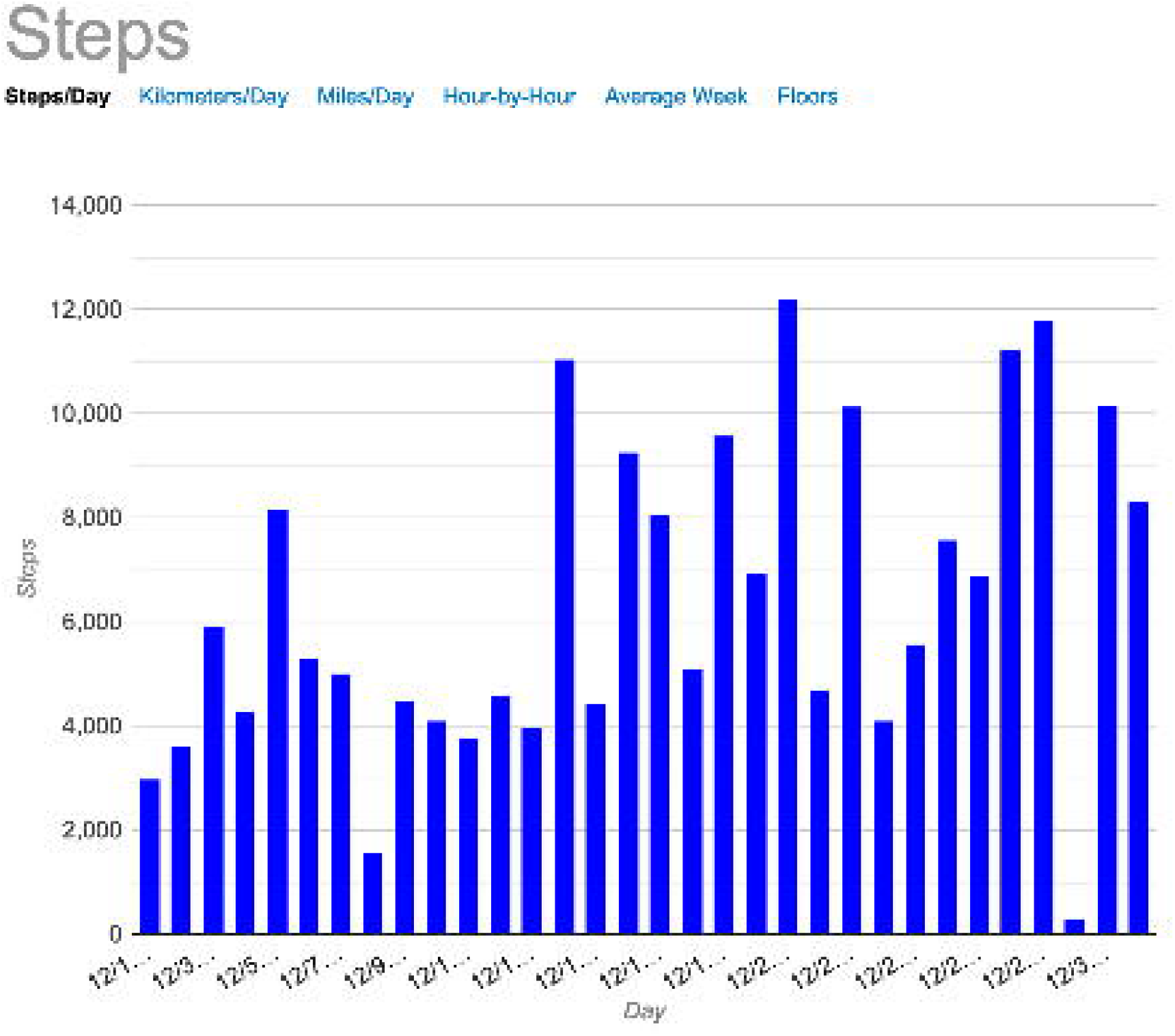
Sample data visualization used in the coaching sessions to review goal attainment, discuss barriers, and set future goals (From the Fitabase platform).

### Participants

Thirteen participants with PD and fourteen participants with HD were enrolled in the *PreActive* intervention. Individuals with PD were recruited through local clinics and support groups and were included if they were 18-85 years old, had a clinical diagnosis of PD Hoehn & Yahr stages I or II, and were able to ambulate independently without an assistive device. Most participants with HD were enrolled through a large clinical trial platform and observational study, Enroll-HD.^40^ Individuals with HD were included if they were 18-65 years old, either premanifest (Unified Huntington Disease Rating Scale^41,42^ Diagnostic Confidence score of 0-3) or in early disease stages (Unified Huntington Disease Rating Scale^41,42^ Total Functional Capacity score of 10-13), and had a minimum score of 24 on the Mini-mental State Examination (MMSE)^43^. All participants were screened using the Physical Activity Readiness Questionnaire (PAR-Q)^44^ for contraindications for exercise, and those who did not pass the PAR-Q were referred to their physician for further screening. Participants with PD or HD were excluded if they had a musculoskeletal injury that would prevent them from participating in exercise, had other neurological disease or uncontrolled psychiatric disorder or behavioral problem, or were already engaging in structured exercise for at least 30 minutes more than three times per week.

Participants provided written informed consent approved by the Institutional Review Board of Teachers College, Columbia University and the New York State Psychiatric Institute (HD only).

### Baseline evaluation

Basic demographics including age, sex, height, weight, and race/ethnicity were collected. All participants underwent a 6-minute walk test and a Stroop interference test. The 6-minute walk test is a standardized clinical measure that assesses walking capacity and general cardiovascular fitness.^45^ The Stroop interference test presents incongruent color-word matches to assess cognitive abilities including attention and executive function.^46^ Additionally, participants with PD completed the Montreal Cognitive Assessment^47^ and the Brunel Lifestyle Physical Activity Questionnaire^48^. The Montreal Cognitive Assessment is reliable and sensitive to detect cognitive impairments in PD.^47^ The Brunel Lifestyle Physical Activity Questionnaire is a valid and reliable measure that provides insight into participants’ planned and unplanned physical activity behaviors.^48^ Participants with HD underwent the Symbol Digit Modality Test and completed the International Physical Activity Questionnaire (IPAQ). The Symbol Digit Modality Test is a brief cognitive assessment that is sensitive to early cognitive changes in HD.^49^ The IPAQ is a widely-validated measure of self-reported physical activity levels.^50,51^

### Data Analyses

#### Data extraction

Daily step count, daily resting heart rate, minute-level heart rate, and minute-level metabolic equivalent of task (MET) data were exported from Fitabase. Data cleaning and reduction were done in Python. Wear time data were calculated throughout the intervention, whereas step count, resting heart rate, and MET data were analyzed for a valid week during the first 14 days and the last 14 days of the intervention. A valid week was defined as a 7-day period with at least 4 valid wear days that included at least 1 weekend day. Baseline data were derived from the earliest valid week within the first 14 days, and follow-up data were derived from the latest valid week within the last 14 days of the intervention.

#### Wear time

Wear time was determined using the total minutes of valid heart rate data available per day. A wear time of ≥600 minutes (10 hours) was deemed a valid wear day, which is the standard accepted threshold.^52^ From this, average wear time per day and the percentage of valid wear days throughout the intervention duration were calculated. Wear time data was plotted and smoothed using a 30-day moving window.

#### Wear habits

Due to an emergence of two patterns of wear, day wearers (average wear time < 16 hours) and day & night wearers (average wear time ≥ 16 hours) were delineated based on an 8-hour sleep schedule, and these groups were used later to explore the influence of Fitbit wear habits on changes in PA from baseline to follow-up of the intervention.

#### Step count, resting heart rate, and MET*min

Average steps and resting heart rate per day was calculated for baseline and follow-up weeks. MET*min was calculated as the MET values multiplied by the number of minutes that MET value appeared, but only when MET ≥ 3.0.

### Statistical analysis

Descriptive statistics on demographics and baseline assessments were calculated separately on participants with PD and HD. Retention rate was calculated based on the number of participants who completed the intervention. Descriptive statistics were used to summarize wear time, expressed in average wear time as well as the percentage of valid wear day during the intervention. Fitbit metrics including step count, resting heart rate, and MET*min per week were compared between baseline and follow-up using effect sizes (Cohen’s d) and calculated separately for participants with PD and HD. The changes in steps and MET*min/week between day wearers and day & night wearers were compared using Cohen’s d. The thresholds for interpreting effect size were small: |d| ≥ 0.2, medium: |d| ≥ 0.5, and large: |d| ≥ 0.8. We did not perform any inference statistics due to the feasibility and exploratory nature of this study, and chose to use an estimation approach to provide insights on effect sizes and trends.^53^

## Results

### Participants

Participants with HD (n=14) were younger and in earlier disease stages than the participants with PD (n=13) (table 1). Due to scheduling availability for pre and post testing, the mean (SD) intervention period for the participants was 101(18) days.

**Table 1.** Participant characteristics at enrollment.

|  | PD (N=13) | HD (N=14) |
| --- | --- | --- |
| Age (years) | 61.7 ± 9.1 | 44.6 ± 11.4 |
| Sex | 9M, 4F | 7M, 7F |
| Height (cm) | 172.8 ± 11.7 | 170.5 ± 9.9 |
| Weight (kg) | 75.9 ± 15.0 | 80.2 ± 17.6 |
| Race/Ethnicity | White = 11<br>Hispanic = 1<br>Asian = 1 | White = 12<br>Black = 2 |
| Disease stage | H&Y I = 4<br>H&Y II = 9 | Pre-manifest = 9<br>Early stage = 5 |
| 6MWT | 442.96 ± 87.61 | 450.17 ± 103.63* |
| Stroop Test | 40.5 ± 10.72 | 44.20 ± 12.57 |
| Cognitive assessments<br>- MoCA | 26.5 ± 2.9 |  |
| Cognitive assessments<br>– Symbol digit<br>modality (#correct) |  | 42.9 ± 17.6<sup>†</sup> |
| Self-reported PA –<br>Brunel (scores 1-5) | Planned PA: 3.85 ± 0.96<br>Unplanned PA: 2.40 ±<br>0.72 |  |
| Self-reported PA –<br>IPAQ<br>(MET*min/week) |  | 3506.50 ± 3264.84 |

\*N=11 due to missing data while study moved remote; <sup>†</sup>N=10 due to missing data. IPAQ, International Physical Activity Questionnaire; MoCA (range 0-30, <26 mild cognitive impairments); Symbol digit modality normative value: 50.

### Feasibility

We assessed feasibility using retention rate and wear time. Retention rate was 85%, with 3 participants dropping out in the HD group and 1 in the PD group. Reasons for dropout were time constraints (n=2), mood disorders (depression and apathy, n=1), and cognitive impairments (n=1). Participants had a mean (SD) of 92.3 (9.2) % valid wear days (wear time ≥ 10 hours) during the intervention period. Average daily wear time was 18.4 (4.5) hours. Wear time per day varied over the intervention period, with an initial 14-day period of higher fluctuation followed by a relatively steady pattern throughout the rest of the intervention (figure 2). Regardless of diagnosis, participants naturally emerged as either day wearers (n=9, average wear time 13.0 (1.2) hours) or day & night wearers (n=14, average wear time 21.9 (1.0) hours) (figure 2).

**Figure 2.**
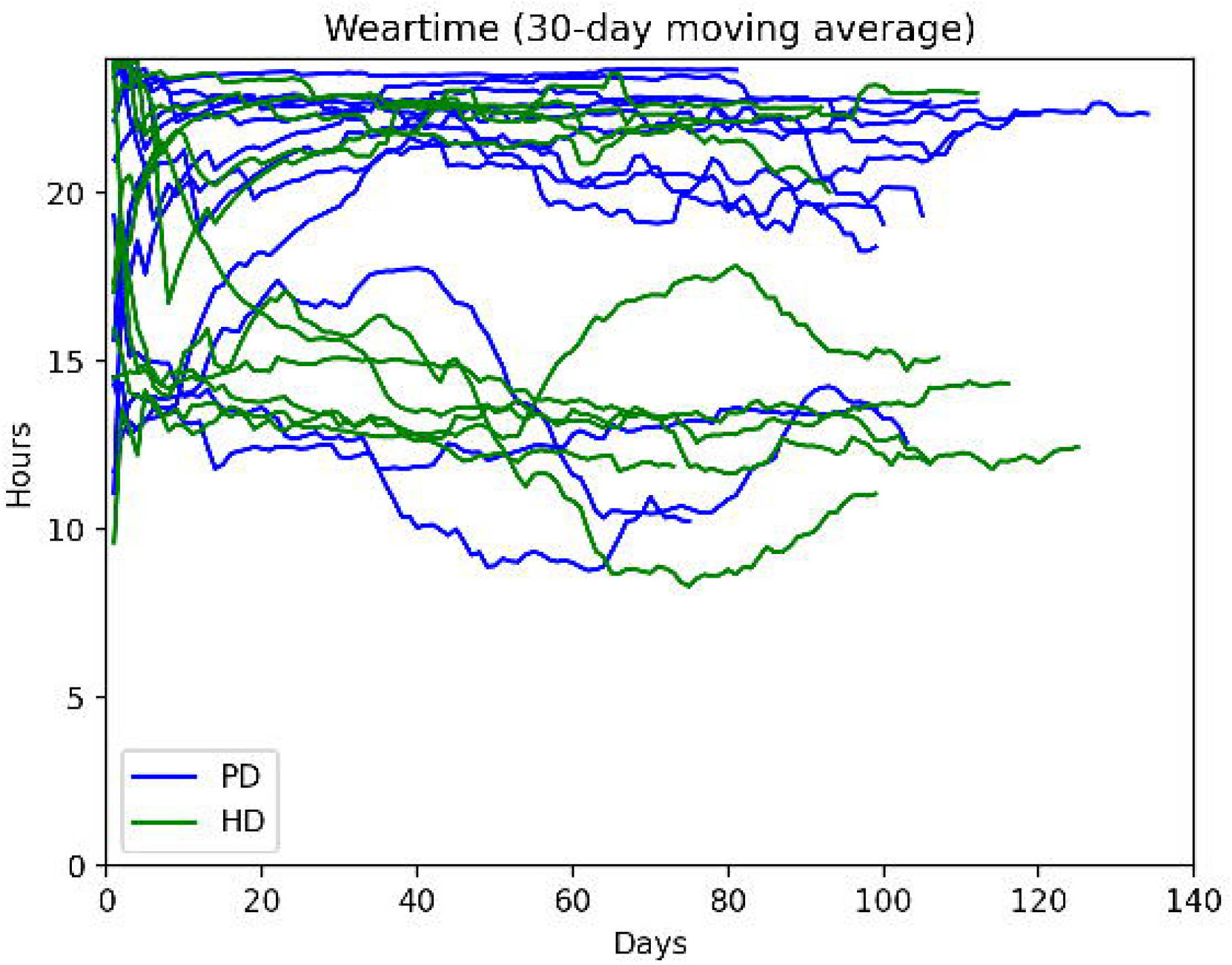
30-day moving average of individual wear time over the course of the intervention. Note an initial period of high fluctuation of wear time before participants settled into two groups representing different Fitbit wear habits: those who were day wearers, and those who were day & night wearers. Key: PD, Parkinson Disease; HD, Huntington Disease.

### Physical activity metrics at baseline and follow-up

There was a small effect of increased steps per day in participants with PD and decreased steps per day in participants with HD from baseline to follow-up (table 2). There was also small effect of increased MET*min per week in participants with PD, but no apparent effect in participants with HD (table 2). There were no apparent effects found in resting heart rate from baseline to follow-up for either participants with PD or HD (table 2).

**Table 2.**
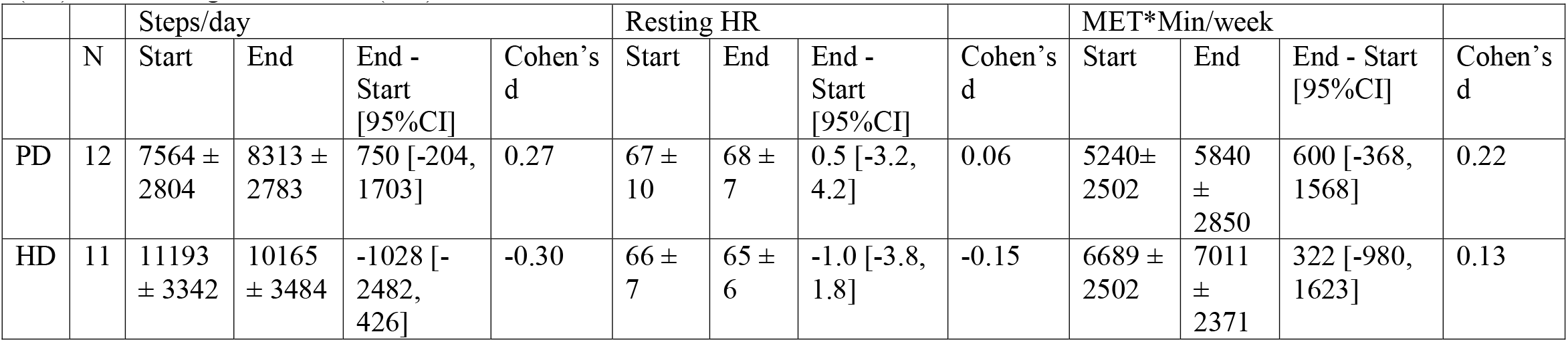
Mean ± standard deviation of Fitbit metrics during qualified week at baseline and follow-up for participants with Parkinson’s (PD) and Huntington’s disease (HD).

### Fitbit wear habits vs. pre-post activity change

We explored whether day wearers and day & night wearers had different changes in steps and MET*min/week from baseline to follow-up (figure 3). Those who were day & night wearers appeared to have greater improvements in steps compared to the day wearers with a large effect size (d=1.02). The day & night wearers also appeared to have greater improvements in MET*min/week compared to the day wearers with a medium effect size (d=0.69).

**Figure 3.**
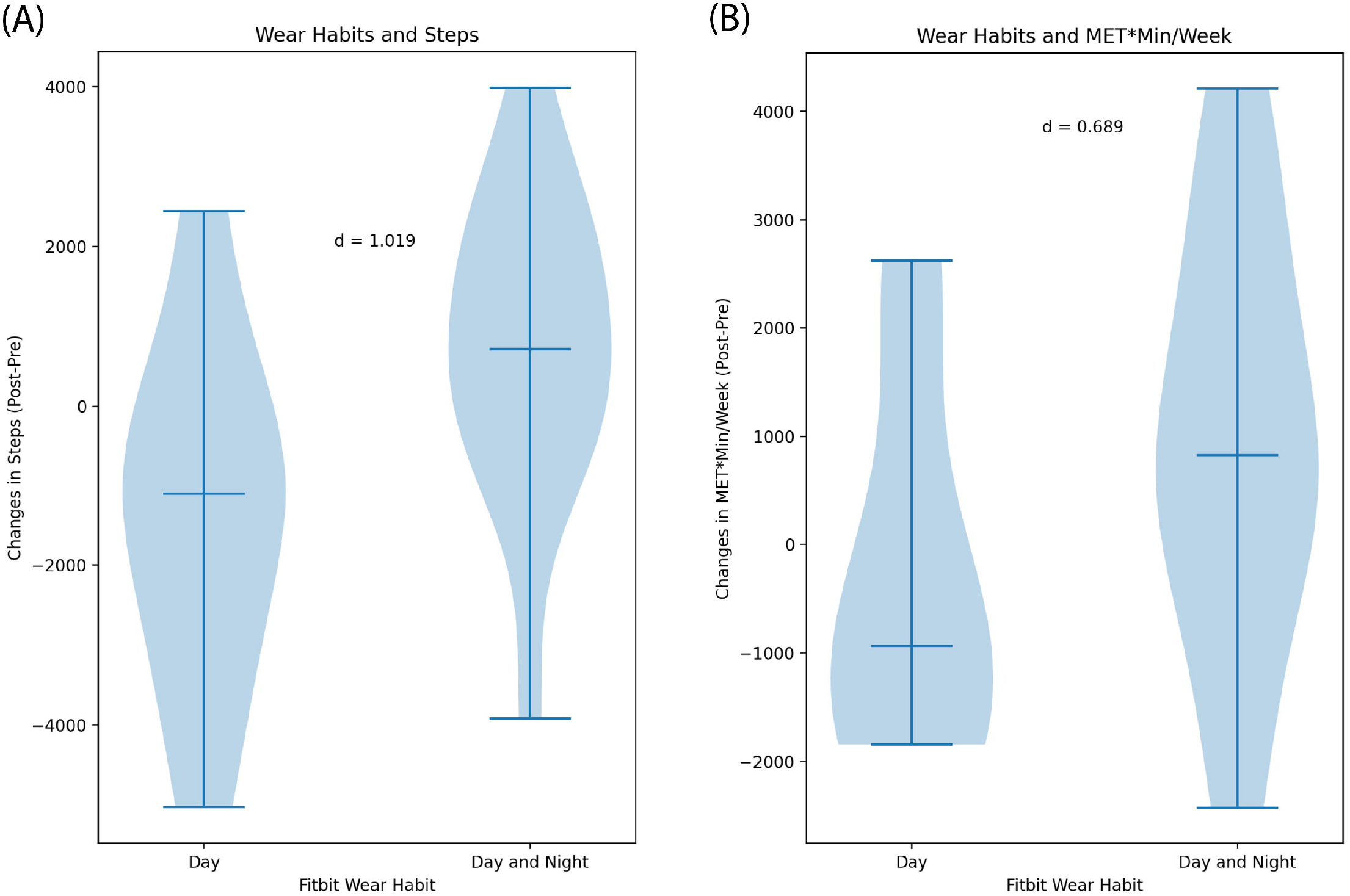
Difference in pre-post changes of (A) steps and (B) MET*min/week between Fitbit day and day & night wearers. Horizontal markers of the violin plots indicate median and interquartile range of the data distribution. The shaded shape of the violin plots indicates the probability density of the data.

## Discussion

This study demonstrated that the use of Fitbits to support therapist-led PA coaching is feasible in people with PD and HD, with high acceptability, retention rate and device wear time. There was a small effect size increase in daily steps and activity (measured as MET*min) in people with PD after the intervention, but not in people with HD. Additionally, those who wore the device day & night showed greater improvements in steps and activity after the intervention compared to those who only wore the device during the day. These findings provide insight into how wearable activity monitors can be implemented in interventions for people with neurodegenerative diseases.

The use of wearable technology holds promise for the monitoring of health behaviors and as a tool to support behavior change.^31,54–56^ This is true across all populations, but especially so for people with PD and HD, who have lower levels of PA and more disease-specific barriers to engage in PA than the general population.^5,26,27^ A device that allows for long-term monitoring of health behaviors could a) identify changes related to disease progression, and b) deliver tailored PA advice. There is a dearth of research on wearables in this population and the current study offers insights on how people with neurodegenerative disease use this technology. Pradhan and Kelly reported high Fitbit adherence in people with mild PD with less than 10% non-wear per day for an average of 13.8 days out of a total trial time of 14 days.^57^ However, long wear time in a short trial like such is not generalizable to longer interventions that span months like *PreActive*. Thus, further research is needed to understand long-term wearable activity monitor use in the neurological population.

The high wear time and percentage of valid wear days (≥10 hrs), showed participant had excellent adherence and tolerated wearing the device well.^58^ When talking with participants, they mentioned that the Fitbit device motivated them, kept them honest about their workouts, provided daily reminders, and encouraged more walking with its step counting function. Most participants also said they checked their data at least daily, indicating high device engagement.

The divide between day wearers and day & night wearers showed that there may be a natural preference of wear habits, perhaps due to comfort during sleep. Moreover, the initial large oscillations of wear time for about 2 weeks may indicate that there is a period of acclimation for the participants to become used to the device and develop their own wearing habits.

Different wear habits that naturally emerged corresponded to different amount of PA uptake throughout the intervention. In fact, more than half of the day & night wearers increased in steps and MET*min, whereas more than half of the day wearers decreased in these metrics (figure 3). Because a decline in PA over time is in line with the natural progression of neurodegenerative diseases,^4^ maintaining or increasing PA during an intervention might be considered a success.

Further to this, the focus of our intervention was to increase time spent in moderate to vigorous intensity PA and not necessarily total daily activity, which may explain the lack of a large increase in step count. We speculate that the differences in PA change between people with different wear habits may be due to the level of commitment and engagement to the device and the *PreActive* program. Even though participants were not instructed to wear the devices to sleep, those who are more enthusiastic and dedicated to improving their health and wellness in general may be more motivated to track their sleep data. It is also possible that activity levels are linked to sleep quality. For example, people with poor sleep (therefore less tolerant wearing a device at night) might find it difficult to increase their activity levels during the day.

There is limited research on the validity of wearable devices in the collection of movement and physiological data on people with PD and HD. de Carvalho Lana et al (2021) found a high correlation (R=.82) in step counts between the Fitbit Charge and the criterion measure (researcher count),^59^ and Keren et al (2021) found similar positive findings among a free-living HD population.^60^ However, given changes in motor symptoms associated with PD and HD, and that physiological responses to exercise may be different in people with PD and HD versus healthy individuals, additional research on validity and reliability of wearables in this population is warranted.

The differences in steps and MET*min between participants with PD and HD at the start of our intervention are potentially revealing distinctions in actual participant behavior and/or bias in measurements. The higher steps and activity measured in those with HD may be due to their younger age and greater agitation and restlessness as reported in some literature,^61^ compared to PD. Given that our participants with HD are all early disease-stage, it is less likely that they have chorea (involuntary movements of the arms), which has been shown to lead to a greater amount of step counts in accelerometry.^60^ One study demonstrated that individuals with early-stage HD have been shown to have higher energy expenditure, which appears to be due to increased physical activity, both voluntary and involuntary.^62^ In our study, we noticed a higher-than-typical MET*min per week calculated from Fitbit data in both participants with PD and HD. For example, MET*min per week data at the start of the intervention was almost double that collected via self-reported IPAQ at baseline for people with HD (Tables 1 & 2). Fitbit does not release information on how their MET estimates are calculated, but indicates that it is estimated based on heart rate for non-step activities, and presumably based on steps for stepping activities.^63^ Furthermore, Fitbit’s “activity minutes” metric, based on >3 METs for more than 10 minutes bouts, has been validated against research-grade accelerometers’ estimate of moderate to vigorous PA.^64^ Assuming Fitbit MET scores are valid METs, the increased MET*min numbers compared to self-report estimates may be due to the passive nature of Fitbit data collection, enabling more activity to be captured that would not have been documented through recall.

There are several limitations to this study. Participants were not blinded to their device data during these weeks used for comparison, so there may have been reactivity, especially in the initial week. Similarly, it is possible that data at the start of the intervention are inflated due the initiation of therapist-led coaching. Due to the variable individual cardiovascular conditions in neurodegenerative diseases, use of age-predicted algorithms is likely not to have been as accurate as maximal exercise testing would have been to inform heart rate response and better define individual’s exercising heart rate zones. However, exercise testing also presents inherent limitations, as tests can be limited by local muscular fatigue or coordination issues with people with PD and HD, thus underestimating maximum heart rate. Another limitation is related to wear time calculation. We determined wear time by the availability of minute-level heart rate data, but it is possible that wear time was underestimated if the device was worn too loose on the wrist, missed measurements, or if the device discarded minute level data after prolonged period of failure to sync. However, if this was the case, actual wear time in this cohort would have been even higher, further supporting the high adherence and feasibility of Fitbit implementation. Finally, we did not systematically collect qualitative data on participant’s experience with Fitbit, and further research understanding their perspective would be invaluable.

Despite wearable activity monitors’ popularity among the general public and in PA research, there are inherent logistical challenges to incorporating them into a study. A recent paper by Balbim and colleagues provided a summary of these challenges and proposed solutions for implementing wearable activity monitors in behavior change intervention studies.^65^ Our experience elicited additional considerations into specific barriers to implement Fitbit in the neurodegenerative population, presented below (table 3).

**Table 3.** Implementation barriers and proposed strategies to using wearable devices in a neurodegenerative population.

| Barriers | Potential Strategies |
| --- | --- |
| Cognitive impairments leading to difficulty learning, wearing, synching, and charging the device. | - Identify care partners available for assistance.<br>- Send scheduled text reminders for synching and charging.<br>Monitor participant data and reach out to remind participants if non-wear or no synching/charging occurred.<br>- Encourage participants to contact research personnel for assistance if they have issues without waiting until next coaching session. |
| Medication or disease affects heart rate response (e.g. beta-blockers). | - Document participants who are taking medications that affects cardiac response to exercise.<br>- Set individual maximum heart rate on the device using results of exercise testing or validated formula for prediction of maximum heart rate in patients on beta-blockers. |
| Neurological symptoms (gait abnormality, tremor, chorea) that affects accelerometry. | - Wear device on the less affected side.<br>- Perform stride length calibration during Fitbit set up.<br>- Collect tremor or chorea symptom severity to account for potential artificial increase in step counts.<br>- Collect perceived physical activity levels using validated questionnaires to cross validate accelerometry. |

Some additional considerations may be valuable for future implementation of wearable activity monitors in PA intervention programs. Blinding device data to reduce reactivity will likely offer a more accurate portrayal of usual PA behaviors. Commercial wearable devices are also intervention tools due to the push notifications and may be hard to blind. Therefore in certain situations that require blinding of data, employing research grade accelerometers should be considered. Use of a data aggregation platform such as Fitabase was found to be very useful for data visualization and management, however, alternative options exist. For example, researchers with sufficient technical knowhow can connect directly to the Fitbit Application Programming Interface (API) to access data. Alternatively, it is viable on a small scale to use individual Fitbit account dashboards for research or clinical programs that have limited funding. To make intervention programs more pragmatic, one may consider allowing participants to bring their own wearable device. If participants are new to wearable activity monitors, it would be valuable to include a 2-week acclimation period whether providing as an intervention and/or using as a measurement tool. Additionally, if sleep data are of interest, considering participants’ willingness to wear the device at night may be an important inclusion criterion. Finally, our coaching program was led by therapists that could guide participants on optimal use of Fitbit monitors, including personalizing goal setting and notifications. We felt this coaching process was important to allow participants to take full advantage of the wearable device and build autonomy for long-term use.

Beyond just tracking activity, wearable monitors have the potential to motivate users through a variety of embedded behavior change techniques such as goal-setting and self-monitoring. In checking and visualizing their own data, controlling when they receive feedback, and deciding how they would like to use this information, users can build autonomy around their health management. By setting and achieving goals based on available metrics from the activity monitors, users can also develop a sense of competence. People with neurodegenerative diseases and other chronic conditions may be more motivated to maintain Fitbit use due to the potential benefit of disease-management.^66^ Importantly, considerations to disease-specific symptoms such as motor, cognitive, and mood impairments must be made when implementing wearable activity monitors in this population. Building sustained PA motivation is a long, but necessary path for people with neurodegenerative diseases to harness this powerful tool for long-term disease self-management. When paired with therapists’ guidance, wearable activity monitors can support each user’s personal journey towards sustained healthy PA practice.

## Data Availability

All data produced in the present study are available upon reasonable request to the authors.

## Acknowledgements

None.

